# Sero-surveillance for IgG to SARS-CoV-2 at antenatal care clinics in two Kenyan referral hospitals

**DOI:** 10.1101/2021.02.05.21250735

**Authors:** R. Lucinde, D. Mugo, C. Bottomley, R Aziza, J. Gitonga, H. Karanja, J. Nyagwange, J. Tuju, P. Wanjiku, E. Nzomo, E. Kamuri, K. Thuranira, S. Agunda, G. Nyutu, A. Etyang, I. M. O. Adetifa, E. Kagucia, S. Uyoga, M. Otiende, E. Otieno, L. Ndwiga, C. N. Agoti, R. Aman, M. Mwangangi, P. Amoth, K. Kasera, A. Nyaguara, W. Ng’ang’a, L. B. Ochola, E. Barasa, P. Bejon, B. Tsofa, L. I. Ochola-Oyier, G. M. Warimwe, A. Agweyu, J. A. G. Scott, K. E. Gallagher

## Abstract

The high proportion of SARS-CoV-2 infections that remain undetected presents a challenge to tracking the progress of the pandemic and implementing control measures in Kenya. We determined the prevalence of IgG to SARS-CoV-2 in residual blood samples from mothers attending antenatal care services at 2 referral hospitals in Kenya. We used a validated IgG ELISA for SARS-Cov-2 spike protein and adjusted the results for assay sensitivity and specificity. In Kenyatta National Hospital, Nairobi, seroprevalence in August 2020 was 49.9% (95% CI 42.7-58.0). In Kilifi County Hospital seroprevalence increased from 1.3% (95% CI 0.04-4.7) in September to 11.0% (95% CI 6.2-16.7) in November 2020. There has been substantial, unobserved transmission of SARS-CoV-2 in parts of Nairobi and Kilifi Counties.

## Background

The proportion of confirmed SARS-CoV-2 infections that have been reported as asymptomatic ranges from 36%-49%^1-4^; in India 94% of a random population sample who had detectable antibodies to SARS-CoV-2 reported having had no symptoms^5^. Symptoms are more common with increasing age and therefore the proportion of infections that are asymptomatic differs with the age-structure of each population. In Kenya, with just 3.9% of the population over 65 years of age^6^, the proportion of infections that are asymptomatic is likely to be high^7^. Additionally, limited access to tests and limited testing capacity makes it likely that a substantial proportion of cases with mild symptoms are not detected. This makes it difficult for the government to track the progress of the pandemic and institute control measures, as symptom based case-finding is likely to miss a substantial proportion of the infections^2^. Measuring the prevalence of antibodies to SARS-CoV-2 is an alternative way to estimate cumulative infection. A number of serological assays have been developed and perform well with high sensitivity and specificity^8-11^. We have shown that 5.6% of blood donors in Kenya had SARS-CoV-2 antibodies in April-June of 2020^12^. However, it is unclear whether blood donors are representative of the population.

In the context of a pandemic, sentinel public health surveillance using residual aliquots of routinely collected blood samples has the potential to overcome participation bias given that unlinked, anonymised residual samples can be tested without informed consent. For example, sero-surveillance for HIV among mothers attending antenatal care was used to track the progress of the HIV pandemic and showed prevalence estimates that were similar to population samples from the same areas^13,14^. In high income countries, a number of seroprevalence studies have indicated that the proportion of pregnant women who have been infected by SARS-CoV-2 is much greater than the cumulative prevalence identified by PCR testing of symptomatic individuals; seroprevalence estimates range from 5-20% but samples differ in time since the virus first emerged^15-21^. It remains unclear whether pregnancy alters susceptibility to SARS-CoV-2 infection^22^; however, a large systematic review has found no difference in risk of becoming symptomatic when comparing pregnant women with confirmed SARS-CoV-2 infection with women of the same age^23^.

In Kenya, in 2014, 50% of women had had at least one pregnancy or were pregnant by 20 years of age and the coverage of at least one antenatal care visit was 96%^24^. Residual blood samples from mothers visiting antenatal care for the first time may represent therefore a relatively unbiased sample of young women, and an alternative to blood donors. Testing an aliquot of blood for antibodies to SARS-CoV-2 is feasible as a venous blood sample (5ml) is already taken to screen mothers for malaria, HIV and syphilis at their first ANC visit. We aimed to determine the prevalence of antibodies against SARS-CoV-2 in mothers attending ANC at 2 referral hospitals in Kenya.

## Methods

### Setting

In a collaboration between the Kenyan Ministry of Health (MOH) and KEMRI-Wellcome Trust Research Programme (KWTRP), two referral hospitals were engaged to provide residual blood samples and limited anonymised data from laboratory register books. Kenyatta National Hospital (KNH) is the national referral tertiary hospital located in Nairobi, the country’s capital city, approximately 3km from the central business district. The population of Nairobi city was 4,397,073 in 2019, the wider urban area includes over 9 million residents^6^. Kilifi County Hospital (KCH) is the county referral hospital in Kilifi Town. The county covers an area of 12,000 km^2^, with a predominantly rural population of 1.4 million^6^. The first SARS-CoV-2 infection in Kenya was identified in Nairobi on the 12^th^ March 2020 and the first infection in Kilifi County was confirmed on the 17^th^ March 2020. By the end of December 2020, 96,458 confirmed cases had been reported in Kenya including 1,670 deaths.

### Study population

All mothers attending ANC for the first time, who provided a routine blood sample, were included in the study. Mothers who did not provide a sample at their first antenatal care visit, or mothers attending their second or subsequent ANC visit, were excluded.

### Sample collection and processing

In Kenya, a 5ml blood sample is routinely collected at the first ANC visit. After testing for malaria, syphilis and HIV in the hospital laboratory, the residual volume is usually discarded. In this study, all residual samples were set aside and collected daily for SARS-CoV-2 sero-surveillance. Where possible, the following data were collected from hospital records and linked to the residual sample identity number: date of sample collection, age, sub-county of residence, trimester of pregnancy and presence or absence of COVID19-like symptoms in the last month. No personal identifiers were collected. In Kilifi, samples were centrifuged, separated and stored at -20 degrees Celsius on the day of collection at the KWTRP laboratories. Samples from Nairobi were processed in a similar manner at the Institute of Primate Research and transported to KWTRP in batches on dry ice. All samples were tested at the KWTRP laboratories for IgG to SARS-CoV-2 whole spike protein using an adaptation of the Krammer Enzyme Linked Immunosorbent Assay (ELISA)^8^. Validation of this assay is described in detail elsewhere^12^. Results were expressed as the ratio of test OD to the OD of the plate negative control; samples with OD ratios greater than two were considered positive for SARS-CoV-2 IgG. Sensitivity, estimated in 174 PCR positive Kenyan adults and a panel of 5 sera from the National Institute of Biological Standards in the UK was 92.7% (95% CI 87.9-96.1%); specificity, estimated in 910 serum samples from Kilifi drawn in 2018 was 99.0% (95% CI 98.1-99.5%)^12^.

### Analysis

We estimated the proportion of samples seropositive for IgG to SARS-CoV-2 by date, location, maternal age, trimester and the presence or absence of symptoms in the last month, as available. Sampling at least 135 mothers per month from each hospital would provide estimates of seroprevalence in the range 3-25% with a precision of 3-7%. Bayesian modelling was used to adjust seroprevalence estimates for the sensitivity and specificity of the assay. Non-informative priors were used for each parameter (sensitivity, specificity and proportion true positive) and the models were fitted using the RStan software package^25^ (see appendix for code). We also obtained the number of PCR-confirmed SARS-CoV-2 infections in the same Counties of residence from the MOH. Sub-county population densities were extracted from the Kenya National Bureau of Statistics’ database^6^.

The study was conducted as anonymous public health surveillance at the request of the MOH. The protocol was also approved by the Scientific and Ethics Review Unit (SERU) of the Kenya Medical Research Institute (Protocol SSC 4085), the Kenyatta National Hospital – University of Nairobi Ethics Review Committee (Protocol P327/06/2020) and the Kilifi County health management team rapid response team.

## Results

### Crude seroprevalence across time and location

Between 30^th^ July and 25^th^ August, 196 samples were collected from Kenyatta National Hospital in Nairobi. Data on age, trimester, sub-county of residence and the presence or absence of COVID19-like symptoms in the month before the visit were available. Women were aged between 17 and 45 years (mean 30 years); 114 (58%) attended their first antenatal care visit during their third trimester of pregnancy. A total of 176 (90%) reported residence in 14 different sub-counties of Nairobi, 64 (35%) of mothers were resident in Embakasi North, East or West sub-counties, and 35 (19%) were resident in Dagoretti North or South sub-counties. Crude seroprevalence was 46% and this did not differ by age, trimester, or the population density in their sub-county of residence. Among 184 (94%) women who had data on symptoms during the preceding month, 12 (7%) reported symptoms and all of those with symptoms reported abdominal pain, which is non-specific for COVID-19. Symptoms were not associated with seropositivity, controlling for age (OR 1.47 95% CI 0.43-5.00; Table 1).

**Table 1.** Seroprevalence of IgG to SARS-CoV-2 among mothers attending antenatal care, by selected characteristics.

|  |  | Total<br>N | Seropositive<br>n | % | Adjusted seroprevalence<br>% | (95% CI) |
| --- | --- | --- | --- | --- | --- | --- |
| **Kenyatta National Hospital, Nairobi** |  |  |  |  |  |  |
| Period (2020) | 30<sup>th</sup> July – 25<sup>th</sup> Aug. | 196 | 91 | 46.4 | 49.9 | 42.7-58.0 |
| Age | 17-29 years | 91 | 38 | 41.8 | 44.7 | 34.2-56.5 |
|  | 30-45 years | 90 | 44 | 48.9 | 52.3 | 40.8-63.7 |
| Trimester | 1 | 17 | 7 | 41.2 | 44.9 | 21.8-70.6 |
|  | 2 | 53 | 21 | 39.6 | 42.8 | 29.1-56.5 |
|  | 3 | 114 | 58 | 50.9 | 54.6 | 44.0-64.9 |
| Any symptoms in last month<sup>1</sup> | Yes | 12 | 7 | 58.3 | 61.2 | 33.2-86.4 |
|  | No | 172 | 78 | 45.4 | 48.7 | 40.4-57.0 |
| Population density of sub-county of residence | <20,000 per km<sup>2</sup> | 97 | 44 | 45.4 | 48.7 | 38.4-59.9 |
|  | 20,000-81,000 per km<sup>2</sup> | 79 | 39 | 49.4 | 52.9 | 40.5-65.0 |
| **Kilifi County Hospital, Kilifi<sup>2</sup>** |  |  |  |  |  |  |
| Month (2020) | September (18<sup>th</sup>-30<sup>th</sup>) | 82 | 0 | 0.0 | 1.3 | 0.04-4.7 |
|  | October (1-31<sup>st</sup>) | 183 | 3 | 1.6 | 1.5 | 0.07-4.1 |
|  | November (1-24<sup>th</sup>) | 154 | 16 | 10.4 | 11.0 | 6.2-16.7 |

<sup>1</sup> Women were asked about the full list of COVID-19 symptoms as per the MOH COVID-19 screening form i.e. fever/ chills, general weakness, cough, sore throat, runny nose, shortness of breath, diarrhoea, nausea/ vomiting, headache, irritability/ confusion, pain (muscular/ chest/ abdominal/ joint).
<sup>2</sup> No age, trimester or symptom data were available from the ANC records at KCH.

At Kilifi County Hospital, 419 samples were collected between the 18^th^ September and 24^th^ November 2020. No data were available on age, trimester, location or symptoms. Crude seroprevalence increased over the period of sample collection from 0% in September to 10% in November (p=0.0001, Chi sq test for trend).

### Adjusted seroprevalence estimates

Seroprevalence, adjusted for assay sensitivity and specificity of the ELISA, was 49.9% (95% CI 42.7-58.0) in Nairobi in August. In Kilifi, it was 1.3% (95% CI 0.04-4.7) and 11.0% (95% CI 6.2-16.7) in September and November, respectively (Table 1).

## Discussion

Surveillance for IgG antibodies to SARS-CoV-2 among mothers attending ANC services in two county referral hospitals in Kenya has revealed evidence of a substantial amount of prior infection in mid-late 2020. Surveillance of pregnant women has been used as a proxy for population-based SARS-CoV-2 surveillance in high income countries^15-21^. This indicates much higher seroprevalence in Kenya than previously reported in samples from blood transfusion donors in the same counties earlier in 2020^12^.

In Nairobi, just after the peak of the first wave of SARS-CoV-2 infections (median date of collection 11^th^ August 2020), 50% of expectant mothers had IgG antibodies to SARS-CoV-2. At the same timepoint, just 6,727 PCR-confirmed infections had been registered across the city (<1% of the County’s population; Figure 1). This estimate is substantially higher than the 7.1% seroprevalence reported among blood donors in the same county by the end of July 2020^12^. We hypothesise the difference between these data and the blood donor data is likely to be due to the localised nature of outbreaks and heterogeneity in the use of KNH across the city. Expectant mothers attending ANC in KNH predominantly came from 5 sub-counties close to the hospital which are densely populated with low-income earners^6^. Blood transfusion donors are likely to be more heterogenous and widely distributed across Nairobi including areas of lower population density and greater affluence.

**Figure 1.**
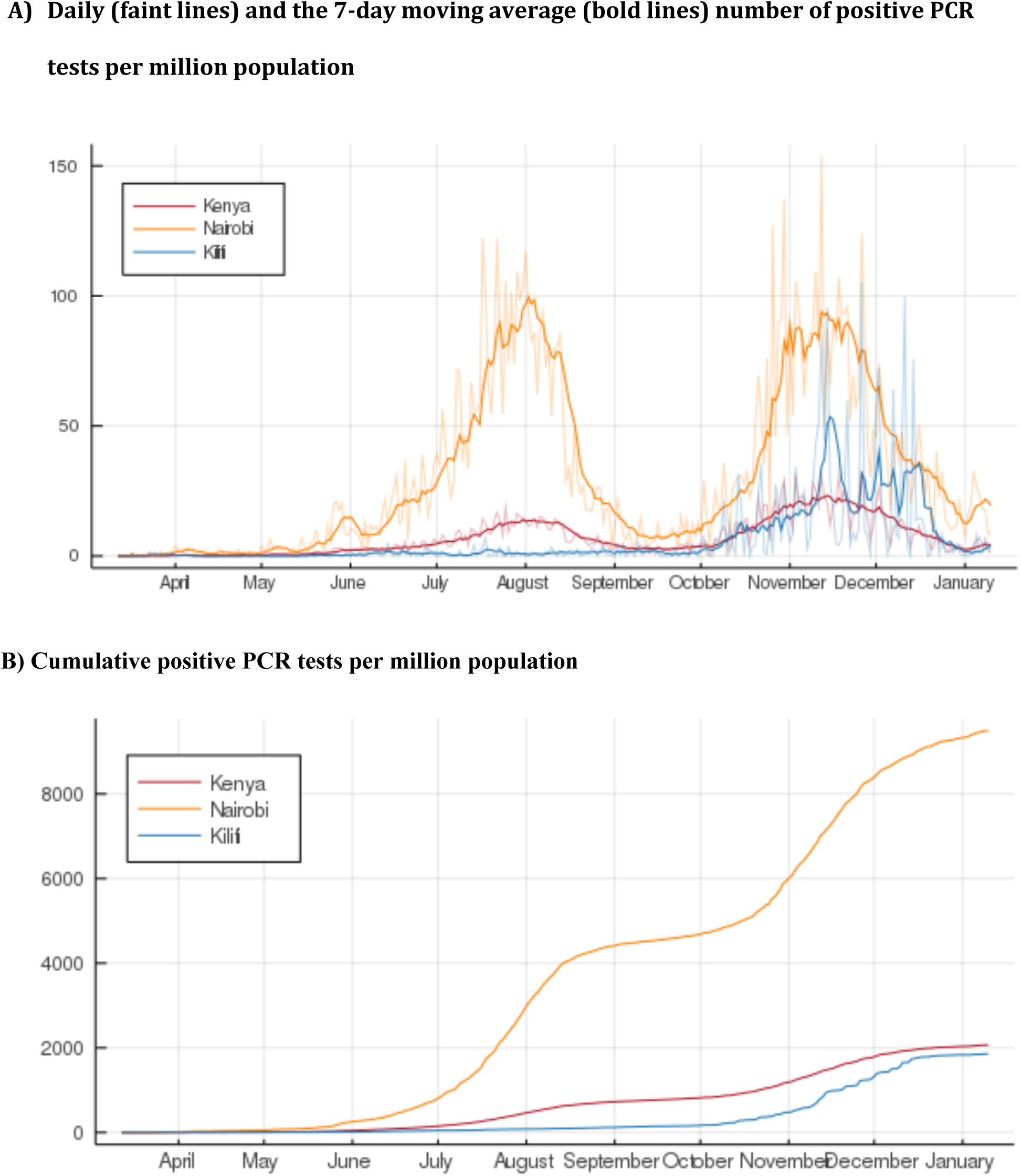
A) Daily number and B) Cumulative daily number of PCR positive tests for SARS-CoV-2 in Kenya, Nairobi and Kilifi Counties.

There was evidence of substantially less prior infection with SARS-CoV-2 among expectant mothers attending ANC in Kilifi County Hospital compared to KNH, but also evidence that seroprevalence had increased markedly during the sampling timeframe to 10.9% by the end of November 2020. The estimate in September (1.3%) was lower than the 4.6% seroprevalence reported among blood donors from Kilifi County (median date of sample collection 30^th^ May)^12^. Again, there is likely substantial geographical heterogeneity in the samples. Blood donations come from across the county including urban centres such as Malindi, whereas women attending ANC are from the hospital catchment area and may represent a less heterogenous group, exposed to less transmission at this timepoint. The lower seroprevalences in Kilifi are consistent with modelling suggesting that the initial wave of the COVID-19 pandemic was concentrated in urban centres, with subsequent spread increasingly affecting rural areas^26^; Kilifi County reported a marked increase in the number of infections in December 2020 (Figure 1).

The serological assay used in this study has been rigorously validated using locally relevant control populations and reference panels from the National Institute for Biological Standards and Control (NIBSC) in the UK^27^. The threshold used to define seropositivity was chosen to prioritise specificity over sensitivity, i.e. to minimize the number of false positives. The very low crude seroprevalence (0%) in the first month of samples from Kilifi adds confirmation that the specificity of this assay is very high.

The impact of pregnancy on susceptibility to SARS-CoV-2 infection is unclear^22^; however, comparisons of infected pregnant women with non-pregnant women of the same age suggests that a similar proportion of infections become symptomatic^23^. This would suggest a similar proportion of pregnant and non-pregnant women mount a protective antibody response and seroprevalence estimates are generalisable to non-pregnant women of the same age. In a comparison of samples from blood donors and ANC in Australia, the two sample sets estimated seroprevalence within 0.1% of each other, although overall prevalence was very low (<1%)^28^. Additionally, seroprevalence in blood donors in Kenya did not differ by sex^12^, suggesting that these results from pregnant women may be generalisable to men between 17-45 years of age, residing in the same areas.

Our analysis is constrained by the nature of the anonymised surveillance data available. Data on age, trimester and location data for the expectant mothers in Kilifi would have allowed more valid comparisons with data from other sources, e.g. blood transfusion donors. The results from Nairobi indicate very low prevalence of COVID-like symptoms even among those who were seropositive; however, stigma associated with disease, particularly early in the pandemic could have resulted in under-reporting. We also lack local data on the rate of antibody waning, which is important to estimate cumulative incidence of infection from snapshot seroprevalence estimates^29,30^. This is especially important in populations, like those reported here, where ongoing transmission may cause ‘natural boosting’ and where circulation of other human coronaviruses may modify immunological responses to SARS-CoV-2^29,31,32^.

## Conclusions

This seroprevalence study of pregnant mothers attending ANC clinics suggests there has been substantial, unobserved transmission of SARS-CoV-2 in parts of Nairobi and Kilifi Counties. Only 7% of women reported any COVID-19-related symptoms, the majority of infections in young adults seem to be asymptomatic. The results suggest at least half of pregnant women in the catchment area of Kenyatta National Hospital in Nairobi and 10% of women in the catchment area of Kilifi County Hospital had already been infected by SARS-SoV-2 by August and November 2020, respectively.

## Data Availability

Data will be made available on reasonable request to the KWTRP Data Governance Committee

## Acknowledgements

We thank the Kenyatta National Hospital and Kilifi county hospital employees who collected the samples during routine ANC visits and the women themselves for providing samples for routine health screening. We thank Rebeccah Ayako, Evalyne Akinyi and Cedrick Shikoli at the Institute of Primate Research for processing the ANC samples from KNH. We thank F. Krammer for providing the plasmids used to generate the spike protein used in this work. Development of SARS-CoV-2 reagents was partially supported by the NIAID Centres of Excellence for Influenza Research and Surveillance (CEIRS) contract HHSN272201400008C. The COVID-19 convalescent plasma panel (NIBSC 20/118) and research reagent for SARS-CoV-2 Ab (NIBSC 20/130) were obtained from the NIBSC, UK. We also thank the WHO SOLIDARITY II network for sharing of protocols and for facilitating the development and distribution of control reagents. This paper has been published with the permission of the director, Kenya Medical Research Institute.

This project was funded by the Wellcome Trust (grants 220991/Z/20/Z and 203077/Z/16/Z), the Bill and Melinda Gates Foundation (INV-017547), and the Foreign Commonwealth and Development Office (FCDO) through the East Africa Research Fund (EARF/ITT/039) and is part of an integrated programme of SARS-CoV-2 sero-surveillance in Kenya led by KEMRI Wellcome Trust Research Programme. ***For the purpose of Open Access, the author has applied a CC-BY public copyright licence to any author accepted manuscript version arising from this submission***.

A.A. is funded by a DFID/MRC/NIHR/Wellcome Trust Joint Global Health Trials Award (MR/R006083/1), J.A.G.S. is funded by a Wellcome Trust Senior Research Fellowship (214320) and the NIHR Health Protection Research Unit in Immunisation, I.M.O.A. is funded by the United Kingdom’s Medical Research Council and Department For International Development through an African Research Leader Fellowship (MR/S005293/1) and by the NIHR-MPRU at UCL (grant 2268427 LSHTM). G.M.W. is supported by a fellowship from the Oak Foundation. C.N.A. is funded by the DELTAS Africa Initiative [DEL-15-003], and the Foreign, Commonwealth and Development Office and Wellcome (220985/Z/20/Z). S.U. is funded by DELTAS Africa Initiative [DEL-15-003], L.I.O.-O. is funded by a Wellcome Trust Intermediate Fellowship (107568/Z/15/Z). R.A is funded by National Institute for Health Research (NIHR) (project reference 17/63/82) using UK aid from the UK Government to support global health research.

The views expressed in this publication are those of the authors and not necessarily those of the funding agencies

## Statisitical appendix: Stan code and input data for estimating adjusted seroprevalence

~~~
data {
 int N;
 int N_se;
 int N_sp;
 int y;
 int x;
 int z;
}
parameters {
 real<lower=0,upper=1> p;
 real<lower=0,upper=1> se;
 real<lower=0,upper=1> sp;
}
transformed parameters {
 real<lower=0,upper=1> p_obs;
 p_obs = se * p + (1 - sp) * (1 - p);
}
model {
 //priors
 p ∼ beta(1, 1);
 se ∼ beta(1, 1);
 sp ∼ beta(1, 1);
 //likelihood
 y ∼ binomial(N, p_obs);
 x ∼ binomial(N_se, se);
 z ∼ binomial(N_sp, sp);
}
~~~

## Data

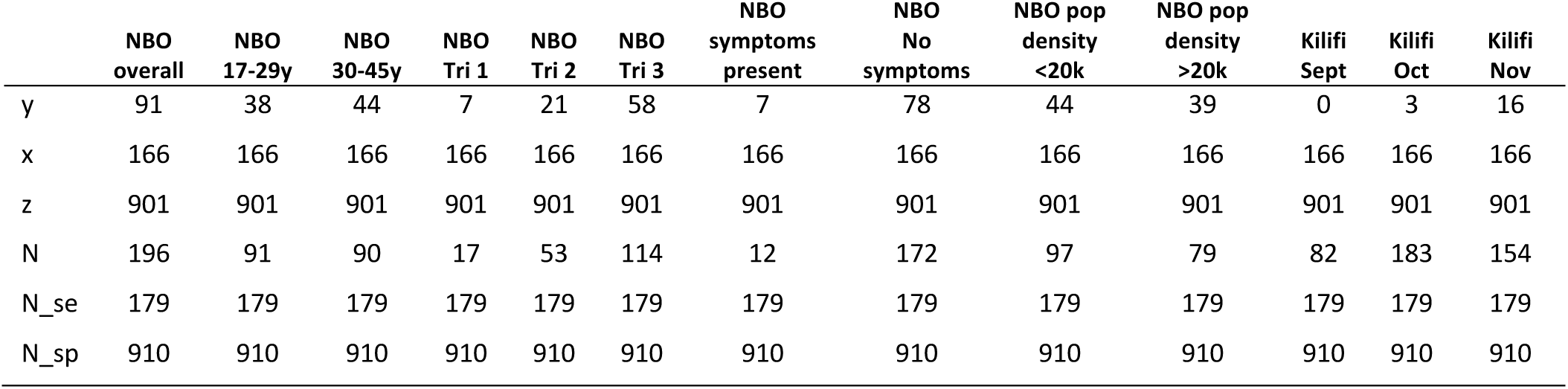

## References

1. Mizumoto K, Kagaya K, Zarebski A, Chowell G. Estimating the asymptomatic proportion of coronavirus disease 2019 (COVID-19) cases on board the Diamond Princess cruise ship, Yokohama, Japan, 2020. Eurosurveillance 2020; 25(10): 2000180.

2. Ng OT, Marimuthu K, Koh V, et al. SARS-CoV-2 seroprevalence and transmission risk factors among high-risk close contacts: a retrospective cohort study. The Lancet Infectious Diseases 2020.

3. Oran DP, Topol EJ. Prevalence of Asymptomatic SARS-CoV-2 Infection. Annals of Internal Medicine 2020; 173(5): 362–7.

4. Al-Qahtani M, AlAli S, AbdulRahman A, Salman Alsayyad A, Otoom S, Atkin SL. The prevalence of asymptomatic and symptomatic COVID-19 in a cohort of quarantined subjects. International Journal of Infectious Diseases 2021; 102: 285–8.

5. Kshatri JS, Bhattacharya D, Kanungo S, et al. Findings from serological surveys (in August 2020) to assess the exposure of adult population to SARS Cov-2 infection in three cities of Odisha, India. medRxiv 2020: 2020.10.11.20210807.

6. Kenya National Bureau of Statistics. Population and Housing Census (www.knbs.or.ke), 2019.

7. Seedat S, Chemaitelly H, Ayoub H, et al. SARS-CoV-2 infection hospitalization, severity, criticality, and fatality rates. medRxiv 2020: 2020.11.29.20240416.

8. Amanat F, Nguyen T, Chromikova V, et al. A serological assay to detect SARS-CoV-2 seroconversion in humans. medRxiv 2020: 2020.03.17.20037713.

9. Lassaunière R, Frische A, Harboe ZB, et al. Evaluation of nine commercial SARS-CoV-2 immunoassays. medRxiv 2020: 2020.04.09.20056325.

10. Whitman JD, Hiatt J, Mowery CT, et al. Test performance evaluation of SARS-CoV-2 serological assays. medRxiv 2020:p 2020.04.25.20074856.

11. Caini S, Bellerba F, Corso F, et al. Meta-analysis of diagnostic performance of serological tests for SARS-CoV-2 antibodies and public health implications. medRxiv 2020:2020.05.03.20084160.

12. Uyoga S, Adetifa IMO, Karanja HK, et al. Seroprevalence of anti–SARS-CoV-2 IgG antibodies in Kenyan blood donors. Science (New York, NY) 2020:eabe1916.

13. Montana LS, Mishra V, Hong R. Comparison of HIV prevalence estimates from antenatal care surveillance and population-based surveys in sub-Saharan Africa. Sexually Transmitted Infections 2008; 84(Suppl 1): i78–i84.

14. Kigadye RM, Klokke A, Nicoll A, et al. Sentinel surveillance for HIV-1 among pregnant women in a developing country: 3 years’ experience and comparison with a population serosurvey. AIDS (London, England) 1993; 7(6): 849–55.

15. Crovetto F, Crispi F, Llurba E, Figueras F, Gomez-Roig MD, Gratacos E. Seroprevalence and clinical spectrum of SARS-CoV-2 infection in the first versus third trimester of pregnancy. medRxiv 2020:2020.06.17.20134098.

16. Mattern J, Vauloup-Fellous C, Zakaria H, et al. Post lockdown COVID-19 seroprevalence and circulation at the time of delivery, France. PLoS One 2020; 15(10): e0240782.

17. Flannery DD, Gouma S, Dhudasia MB, et al. SARS-CoV-2 seroprevalence among parturient women in Philadelphia. Science immunology 2020; 5(49).

18. Cosma S, Borella F, Carosso A, et al. The “scar” of a pandemic: Cumulative incidence of COVID-19 during the first trimester of pregnancy. Journal of medical virology 2020.

19. Villalaín C, Herraiz I, Luczkowiak J, et al. Seroprevalence analysis of SARS-CoV-2 in pregnant women along the first pandemic outbreak and perinatal outcome. PLoS One 2020; 15(11): e0243029.

20. Lumley SF, Eyre DW, McNaughton AL, et al. SARS-CoV-2 antibody prevalence, titres and neutralising activity in an antenatal cohort, United Kingdom, 14 April to 15 June 2020. Eurosurveillance 2020; 25(42).

21. Haizler-Cohen L, Davidov A, Blitz MJ, Fruhman G. Severe acute respiratory syndrome coronavirus 2 antibodies in pregnant women admitted to labor and delivery units. American journal of obstetrics and gynecology 2020.

22. Wastnedge EAN, Reynolds RM, van Boeckel SR, et al. Pregnancy and COVID-19. Physiol Rev 2021; 101(1): 303–18.

23. Allotey J, Stallings E, Bonet M, et al. Clinical manifestations, risk factors, and maternal and perinatal outcomes of coronavirus disease 2019 in pregnancy: living systematic review and meta-analysis. BMJ 2020; 370: m3320.

24. Kenya National Bureau of S, Ministry of HK, National ACCK, Kenya Medical Research I, National Council for P, Development/Kenya. Kenya Demographic and Health Survey 2014. Rockville, MD, USA, 2015.

25. Stan Development Team. RStan: the R interface to Stan. R package version 2.21. 2, http://mc-stan.org/. 2020.

26. Ojal J, Brand SP, Were V, et al. Revealing the extent of the COVID-19 pandemic in Kenya based on serological and PCR-test data. medRxiv 2020:2020.09.02.20186817.

27. Giada Mattiuzzo, Emma M. Bentley, Mark Hassall, et al. Expert Committee on Biological Standardisation: Establishment of the WHO International Standard and Reference Panel for anti-SARS-CoV-2 antibody; Geneva 9–10 December 2020, 2020.

28. Gidding HF, Machalek DA, Hendry AJ, et al. Seroprevalence of SARS-CoV-2-specific antibodies in Sydney, Australia following the first epidemic wave in 2020. Med J Aust 2020; https://www.mja.com.au/journal/2020/seroprevalence-sars-cov-2-specific-antibodies-sydney-australia-following-first [Preprint, 2 November 2020]. 2020.

29. Huang AT, Garcia-Carreras B, Hitchings MDT, et al. A systematic review of antibody mediated immunity to coronaviruses: kinetics, correlates of protection, and association with severity. Nature Communications 2020; 11(1): 4704.

30. Buss LF, Prete CA, Jr., Abrahim CMM, et al. Three-quarters attack rate of SARS-CoV-2 in the Brazilian Amazon during a largely unmitigated epidemic. Science (New York, NY) 2021; 371(6526): 288–92.

31. Le Bert N, Tan AT, Kunasegaran K, et al. SARS-CoV-2-specific T cell immunity in cases of COVID-19 and SARS, and uninfected controls. Nature 2020; 584(7821): 457–62.

32. Otieno G, Murunga N, Agoti C, Gallagher K, Awori J, Nokes D. Surveillance of endemic human coronaviruses (HCoV-NL63, OC43 and 229E) associated with childhood pneumonia in Kilifi, Kenya [version 2; peer review: 2 approved]. Wellcome Open Research 2020; 5(150).

